# Mortality according to gender identity and sexual orientation: Data relationship strategies for Rio de Janeiro, Brazil

**DOI:** 10.1101/2024.01.22.24301609

**Authors:** Ricardo de Mattos Russo Rafael, Kleison Pereira da Silva, Helena Gonçalves de Souza Santos, Davi Gomes Depret, Jaime Alonso Caravaca-Morera, Karen Marie Lucas Breda

**Affiliations:** State University of Rio de Janeiro, School of Nursing, Public Health Nursing Department. Rio de Janeiro, Brazil; University of Costa Rica. San José, Costa Rica; University of Hartford, Department of Nursing, College of Education, Nursing & Health Professions. West Hartford, Connecticut, United States of America

**Keywords:** Gender identity, sexual orientation, probability, mortality records

## Abstract

**Objective:** To evaluate the accuracy, potential, and limits of probabilistic data relationships to yield information on deaths according to sex identity and sexual orientation in the state of Rio de Janeiro.

**Methods:** This study evaluated the accuracy of the probabilistic relationship of data to obtain information on deaths according to gender and sexual orientation. Data from two information systems were used from June 15, 2015 to December 31, 2020. We constructed nine probabilistic data relationship strategies and identified the performance and cutoff points of the best strategy.

**Results:** The best data blocking strategy was established through logical blocks with the first and last names, birthdate, and mother’s name in the pairing strategy. With a population base of 80,178 records, 1556 deaths were retrieved. With an area under the curve of 0.979, this strategy presented 93.26% accuracy, 98.46% sensitivity, and 90.04% specificity for the cutoff point ≥ 17.9 of the data relationship score. The adoption of the cutoff point optimized the manual review phase, identifying 2259 (90.04%) of the 2509 false pairs and identifying 1532 (98.46%) of the 1556 true pairs. A crude mortality rate of 19.11 deaths per 1,000 people was observed, where women who had sex with women, transvestites, and transgender women had higher mortality rates than heterosexual cisgender women. Neither men nor men with gender markers had higher mortality rates than heterosexual cisgender women.

**Conclusion:** With the identification of possible strategies for determining probabilistic data relationships, the retrieval of information on mortality according to sexual and sex markers has become feasible. Based on information from the daily routine of health services, the formulation of public policies that consider the LGBT+ population more closely reflects the reality experienced by these population groups.

## INTRODUCTION

Mortality has been considered an important indicator in the context of public health, especially in the area of health surveillance and for public policymakers in Brazil and worldwide. It is not by chance that numerous studies address this topic (1–5). This is because monitoring of death rates not only helps to identify the patterns and causes of death of population groups but also is useful in defining priorities for the allocation of resources and evaluating the quality of life and well-being of populations, in addition to allowing the measurement of the efficiency of the programs implemented (6,7). Given the importance of monitoring these data, since the 1970s Brazil has implemented one of the most robust information systems for monitoring deaths, the mortality information system (SIM). Under the administration of the Brazilian Ministry of Health and fully integrated into the Unified Health System, SIM can publish detailed data on the cause of death using the International Classification of Diseases-10 and several individual and clinical characteristics (8–10).

On the other hand, it is still not possible to say that information on deaths truly reaches all population groups. Paradoxically, although the literature is vast that points out that gays, lesbians, bisexuals, *travestis*, and transgender people (LGBT+; using the “plus” sign to represent the variety of sexual orientations and gender identities that exist) have worse health indicators than others (11–16), SIM still does not monitor specific information on sexual orientation and gender identity. Thus, *travestis* and trans women who were unable (or did not have the desire to) to rectify their names had their death records classified as belonging to the male population. The same occurs for trans men, who are classified as part of the female population. This is because only the sex assigned at birth and recorded in Brazilian administrative systems is identified in SIM (17,18). This is the paradox of the country that, according to nongovernmental agencies, murders the most LGBT+ people (19).

In the absence of specific information and due to the urgency of providing data that support specific public policies for these groups, the National Association of *Travestis* and Transgenders (ANTRA – *Assoiação Nacional de Travestis e Transexuais*), as well as gay activist groups, monitors deaths based on newspaper reports and information captured from hospital nets (19). These cases, exclusively focused on murders, represent only a small part of the concrete reality experienced by these people. In other words, the country is very far from familiar with the mortality profile of the LGBT+ population, harming everything from the design of public policies to the direct care provided by health professionals (still under the fog of the unknown due to the absence of data).

In all Brazilian health information systems, information on sexual orientation and gender identity has been systematically collected only from the interpersonal and self-inflicted violence module of the Notifiable Diseases Information System (SINAN) since June 15, 2015 (20). All suspected or confirmed violent events are recorded, providing a way to expand the dataset on the LGBT+ population. With possible inaccuracies due to the absence of a unique key to identify the population in information systems, the possibility of a probabilistic relationship of data may suggest a tool to estimate deaths in the Brazilian LGBT+ population. Data relationships are a statistical tool for combining different sets of information: deterministically, when there is a unique key (identifying variable) for linking, or probabilistically when keys are used to estimate the probability that records with different names refer to the same person (21–26). As a way of contributing to the systematic integration of the technique in health services, this study aims to evaluate the accuracy, potential, and limits of the probabilistic relationship of data to obtain information on deaths according to sex and sexual orientation in the state of Rio de Janeiro.

## Methods

### Study type

This study quantifies the accuracy with which the probabilistic relationships of data yield information on deaths according to gender identity and sexual orientation, which integrates the analyses of the first five years of the study "Effects of (trans)gender identity and sexual orientation on notification and mortality due to violence: a cohort study".

### Sources, place, and period of study

This study used data from people registered in the SINAN for interpersonal and self-inflicted violence and from the SIM data of 92 municipalities in the state of Rio de Janeiro, Brazil, from June 15, 2015 to December 31, 2020.

### Population and selection criteria

To determine the probabilistic relationships between the databases, the SINAN records and occurrence of interpersonal/self-inflicted violence between June 15, 2015 and December 31, 2020 and the times of introduction of the variables “sexual orientation” and “gender identity” in the notification form were included. The records of adults aged 19 to 59 years were included. Records without information or with invalid data on birthdate, name, mother’s name, and date of occurrence of violent acts were extracted. For SIM qualification, records without identification or with invalid names or mothers’ names (e.g., indigent, male, black male, white female, etc.) were excluded. To reduce the number of records and accelerate the data relationship process, records of people younger than 19 years of age were excluded, as this was the criterion for selection in SINAN.

### Study variables

#### Variables of the probabilistic relationship of the data

The variables used as link keys between the databases were “name”, “mother’s name”, and “birthdate”. In addition, the variables “notification number”, “date of notification”, “date of occurrence”, “death number”, and “date of death” were used in SINAN to construct an identifier code and to verify duplicate records.

#### Outcome and exhibits of interest

The outcome variable was death (all causes), which was identified and retrieved from SIM through the database link. Variables such as sex, sexual orientation, and gender identity, which were exposures of interest in this investigation, were synthesized to build analytical groups approximating the realities of exposure of people to the concrete reality. Originally present in SINAN, the possible entries included female (S0) and male (S1) for the variable “sex”; heterosexual (O0), bisexual or homosexual (O1), and “unknown” or “omitted” (O2) for “sexual orientation”; and, for the gender variable, it is important to consider that the registration was made only for *travestis* and trans women (G1) and trans men (G2), with the interpretation that data “unknown” or “omitted” (G0) referred to the cisgender population, despite the validity problems that this may produce (18).

Thus, eight groups were created. The participants were cisgender and heterosexual women (S0+O0+G0), women who had sex with women (S0+O1+G0), cisgender women of unknown sexual orientation (S0+O2+G0), *travestis* and trans women (G1), cisgender and heterosexual men (S1+O0+G0), men who had sex with men (S1+O1+G0), cisgender men with unknown sexual orientation (S1+O2+G0), and trans men (G2). Due to the limited number of cases retrieved from the databases, the sexual orientations of transgender people were not combined in the groups formed, limiting the definition of interest groups.

#### Other covariates

The other covariates tested in the study, all derived from SINAN, were “age group” based on the transformation of the continuous numerical variable “age” into four strata of adults (19-29; 30-39; 40-49; ≥50). The exclusion criteria for individuals were as follows: marital status (married, single, and other), time of schooling (up to 8 years, more than 8 years), person with disability (no, yes when at least one disability was recorded), people with mental disorders and behavioral disorders (no, yes when at least one disorder was recorded), and referral to the care network (no yes when at least one referral to social, legal, or health care services was documented). The variable “race/color” was classified as black (black and mixed-race) or nonblack, following an ethical-political and conceptual approach adopted in investigations with this focus in the Brazilian context (28). The variable “chronicity of violent episodes” was created from the repeated insertion of records into SINAN, as detailed in the section on probabilistic data relationships.

### Data processing and analysis

#### Routines of the probabilistic relationship of data

The first stage, titled preprocessing, was intended for the preparation and standardization of the databases for the probabilistic relationship of the data (22). In this stage, computational scripts were built in Python 3.11.2 software in the Visual Studio Code 1.84.2 environment. Initially, the databases were reduced, leaving only the variables of interest for linking the databases (notification number/death number, date of notification, date of occurrence/date of death, name, mother’s name, and birthdate) to facilitate the process. The study selection criteria were also applied, and unique keys were constructed for each record in the database. This construction was necessary because, at least in SINAN, the notification number, which should be unique for each record, shares the same number with other people. In other words, different people had the same notification number without being homonymous. Thus, the unique key was generated based on the combination of the notification (or death) number, birthdate, first and last name, and mother’s name of each person.

With the expectation of reducing potential spelling errors in name records, a set of standardizations was implemented. All letters were converted to uppercase letters. Accents, special characters, excessive spaces, and other punctuation were eliminated (“Ç” became “C”, “Á” became “A”, etc.), as were prepositions (“DE”, “DE”, “ DO”, "DA", "DOS", "DAS", "E"). Duplicate letters were kept only once (e.g., “TT” and “SS” became “T” and “S”, respectively). The letters were standardized according to Portuguese phonetics. In this context, the syllables “WA”, “KA”, “KO”, “KU”, “CE”, “CI”, “GE”, and “GI” were replaced by “VA”, “CA”, “CO”, “CU”, “SE”, “SI”, “JE”, and “JI”, respectively. The names starting with “H” had this letter deleted (e.g., “HUGO” became “UGO”), and the letter “Y” was replaced by “I”. To handle last names in the preprocessing phase, the names were separated into fragments (first name, middle name, and last name) to optimize the next steps of building data relationships. Dates were standardized to write the day, month, and year as two, two, and four numerals, without separation by slashes.

The following steps, which included the completion of database preparation and the steps related to the probabilistic relationship of the data itself, were conducted in Link Plus version 3.0. This software, developed by the USA’s Centers for Disease Control and Prevention, was initially designed for the probabilistic relationship of cancer registries in the USA (29). However, in the field of public health, its use has spread in other contexts and countries, including Brazil (23,26,30–32).

Thus, to finalize the preparation of the databases, the deduplication technique was applied due to the possibility of multiple records in SINAN since the recurrence of different violent events throughout life and multiple notifications of the same events are quite likely. Events may be recorded by more than one professional or health unit. The deduplication technique consists of verifying these repeated records in a database (22,24). Cases of duplicity were treated according to the following rules: 1) When the occurrence number was the same, with the same notification and occurrence dates, victim’s name, mother’s name, and birthdate, even allowing for spelling variations indicative of error, the record was considered the same, so only one such record was selected. 2) When the date of occurrence was different but the name, mother’s name, and birthdate were the same, even with the presence of spelling changes, it was considered a recurrence of the event, which we treated as a recurrent/chronic case. In this second rule, only the oldest record was selected, and we counted the number of repeated events in a new variable named “event chronicity”.

After standardization and preprocessing were completed, the blocking and pairing stage began. Blocking is a method of creating logical subsets based on specific criteria of variables to reduce the number of comparisons during the database relationship. This reduces the processing time. Pairing is intended to compare the corresponding records in the databases using preestablished algorithms in the relationship software (22,24).

In the specific case of this study, nine blocking and pairing strategies were compared. The blocking strategies combined the first name (FN), last name (LN), mother’s first name (FM), mother’s last name (LM), and birthdate (BD) using the soundex code to reduce potential spelling errors and nominal variations. For the pairing strategies, combinations of the full name (N), mother’s name (M), and birthdate (BD) were used, defining the minimum probabilities of agreement (M-probability) as 0.95 and 0.65 for the name and birthdate, respectively. In addition, the direct method of pairing and an initial cutoff point of 10 were used for the scores calculated from the data.

A manual peer review was performed for each strategy. The defining criteria for true pairs were as follows: 1) same name, birthdate, and mother’s name; 2) same name, mother’s name, and day and month of birth but with a variation of ±2 years for the last digit of the year of birth; 3) same name, mother’s name, and year of birth, with inversion of the day and month digits; 4) same name, mother’s name, and day and year of birth; 5) same name, mother’s name, and month and year of birth; 6) similar name and mother’s name with variation in the orthographic scan; and 7) unusual name and mother’s name (foreign names, for example) that agree on the date of birth.

In the postpairing phase, a new deduplication was applied to the sets of pairs formed in each strategy. The selection of the best matching and blocking strategy was based on the shortest processing time and the most true pairs after deduplication. Once the best strategy was found, the files were combined based on the unique key constructed during the preparation phase, thus joining the true pairs (deaths) with the SINAN database and all its variables. The same occurred with the SIM variables, specifically concerning true pairs. The bank combinations were performed using Stata SE 15 software.

#### Statistical analysis

The processing times, minimum and maximum scores, absolute numbers, and proportions of true pairs formed by each blocking and pairing strategy of the probabilistic data relationship were analyzed. The criteria that made up the best strategy had receiver operating characteristic (ROC) curves drawn and the areas under the curves calculated to define the best cutoff point for the matching scores. In addition, the sensitivity (%), specificity (%), accuracy (%), positive and negative likelihood ratios, number of true pairs, and number of false pairs were calculated for each cutoff point of the scores of the chosen strategy.

After the SINAN database was integrated with the true pair data defined by the chosen strategy, the proportions corresponding to each covariate of interest in the study were calculated. This included the number of identified deaths and the crude mortality rates per thousand people, comparing the initial cutoff point with that selected in the most effective matching strategy. In addition, with the objective of observing the existence of differences between the cutoff points, the proportion of agreement, the kappa statistic, and the respective p values were calculated for each covariate of the study. The interpretation of the kappa statistic was classified as full agreement (0.81- 1.00), substantial agreement (0.61-0.80), moderate agreement (0.41-0.60), fair agreement (0.21-0.40), slight agreement (0-0.20), or no agreement (<0) (33,34). All analyses were performed using Stata SE 15 software.

### Ethical aspects

This study followed all the ethical principles of research involving human beings. Because this study was based on information from two databases that were systematically collected by health professionals, there was no need to sign a free and informed consent form. As personal and confidential information was needed, such as names and addresses, the nonanonymized data were manipulated only by the project coordinator and, later, coded and separated into separate files, aiming to maintain the privacy of the information. The research project from which the results of this study emerged was approved by the research ethics committee of the State University of Rio de Janeiro (Rio de Janeiro, Brazil) under protocol number 5,009,244.

## Results

Figure 1 shows the sample composition that formed the baseline of the SINAN probabilistic relationship of interpersonal and self-inflicted violence, as well as the SIM rating. Note that 60.35% (n = 128,620) of the records were excluded from SINAN based on the selection criteria. In addition, 3639 cases of notification recurrence were identified—i.e., when the case was the same but with different events and dates. In this case, only one record was kept, for which purpose we created a specific variable, which represented the chronicity of violent episodes. Thus, the baseline consisted of 80,178 records. In turn, SIM had 90,279 (10.12%) exclusions of records based on the qualification criteria of the database, totaling 737,493 death records (Figure 1).

**Figure 1.**
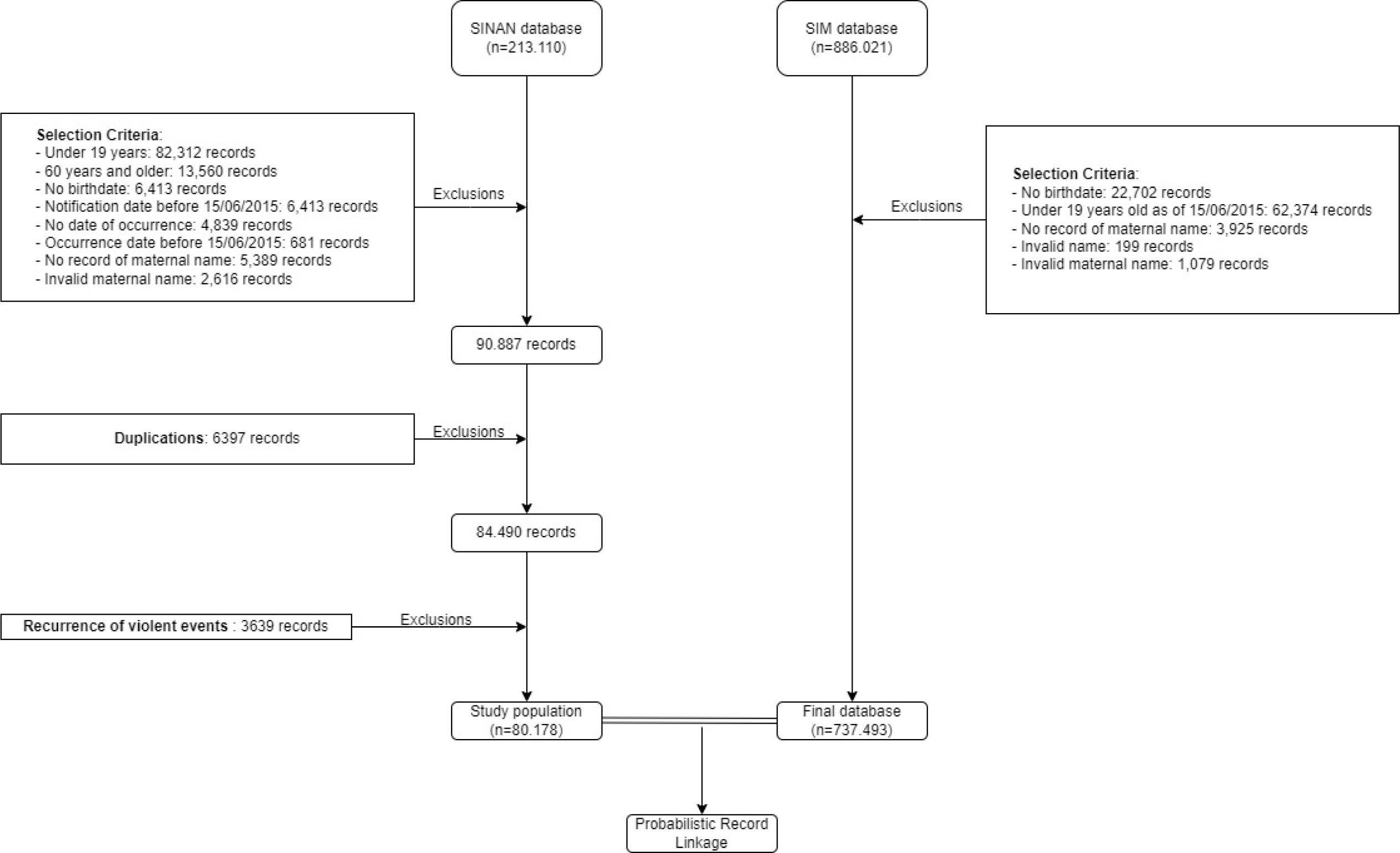
Flowchart of the study population composition for probabilistic record linkage. State of Rio de Janeiro, Brazil, 2015-2020.

**Table 1** presents the results of the probabilistic relationship according to the 9 blocking and pairing strategies. strategy 1, although not the one with the highest percentage of true positives after deduplication, had the highest absolute number of true pairs (n = 1156) and the shortest data processing time (11 minutes and 50 seconds). Therefore, according to the preestablished criteria, the best strategy for further analysis was strategy 1.

**Table 1 .** The results of probabilistic record linkage according to blocking and matching strategies. State of Rio de Janeiro, Brazil, <u>2015-2020.</u>

| ID | Blocking strategy | Matching strategy | Score | Processing Time | P1 | TP1 | %TP1 | PD | P2 | TP2 | %TP2 |
| --- | --- | --- | --- | --- | --- | --- | --- | --- | --- | --- | --- |
| 1 | FN + LM | N + BD + M | 10.6-29.7 | 00:11:50 | 4087 | 1578 | 38.61 | 22 | 4065 | 1556 | 38.28 |
| 2 | FN + LN + BD + FM + LM | N + BD + M | 10.6-29.7 | 02:28:54 | 7296 | 1570 | 21.52 | 19 | 7277 | 1551 | 21.31 |
| 3 | FN + LN + FM + LM | N + BD + M | 10.6-29.7 | 00:25:46 | 7262 | 1570 | 21.62 | 20 | 7242 | 1550 | 21.4 |
| 4 | FN + LN + FM + LM | N + BD | 12.1-20.0 | 00:20:24 | 2388 | 1111 | 46.52 | 10 | 2378 | 1101 | 46.3 |
| 5 | FN + LN + FM + LM | BD + M | 10.6-18.5 | 00:19:11 | 5561 | 996 | 17.91 | 9 | 5552 | 987 | 17.78 |
| 6 | FN + LN + BD | N + BD + M | 10.6-29.7 | 02:19:07 | 7296 | 1570 | 21.52 | 20 | 7276 | 1550 | 21.3 |
| 7 | FN + LN + BD | N + BD | 12.1-20.0 | 01:32:52 | 2397 | 1103 | 46.02 | 10 | 2387 | 1093 | 45.79 |
| 8 | FN + BD | N + BD + M | 10.6-29.7 | 02:10:48 | 7296 | 1570 | 21.52 | 20 | 7276 | 1550 | 21.3 |
| 9 | BD | N + BD + M | 10.6-29.7 | 02:07:21 | 7296 | 1570 | 21.52 | 20 | 7276 | 1550 | 21.3 |

**ID:** identification of the data linkage strategy; **P1:** pairs formed after data linkage; **TP1:** proportion of true pairs formed after data linkage; **DP:** number of duplicate pairs; **P2:** pairs formed after the deduplication strategy; **TP2:** true pairs formed after deduplication; **%TP2:** proportion of true pairs formed after deduplication; **DB:** birthdate; **N:** name; **FN:** soundex of the first name; **LN:** soundex of the last surname; **M:** mother's name; **FM:** soundex of the mother's first name; **LM:** soundex of the mother's last surname.

Figure 2 shows the ROC curve of strategy 1. With an area under the curve of 0.979 (95% confidence interval: 0.976-0.983), the probabilistic relationship has an excellent ability to discriminate between true and false pairs. **Table 2** lists the properties of the strategy. When a cutoff point of 17.9 was used for the score, the sensitivity was 98.46%, the specificity was 90.04%, and the positive and negative likelihood ratios were 9.84 and 0.02, respectively. This means this cutoff point allowed the correct identification of 98.46% of the true pairs and 93.26% of the false pairs. In addition, the likelihood ratio values show that a positive rating in the relationship significantly increased the chances of the pair being truly positive, while a negative classification, due to the very low negative likelihood ratio, provided strong evidence that the pair was not a true pair. Finally, when analyzing the accuracy, we observed that this cutoff point could correctly classify 93.26% of the observations, i.e., 1532 (98.46%) of the 1556 true pairs, as well as correctly classify 2259 (90.04%) of the 2509 false pairs, greatly reducing the need for manual review after the probabilistic relationship of the data.

**Table 2 .** Accuracy by cutoff point of the best blocking and matching strategy (strategy 1). State of Rio de Janeiro, Brazil, 2015-2020 (n=4065)

| **Cutoff** | **S%** | **E%** | **Ac%** | **LR+** | **LR-** | **TP** | **FP** |
| --- | --- | --- | --- | --- | --- | --- | --- |
| ≥ 10.6 | 100.00 | - | 38.28 | - | - | 0 | 906 |
| ≥ 12.1 | 100.00 | 36.11 | 60.57 | 1.00 | - | 3 | 1.088 |
| ≥ 12.3 | 99.81 | 79.47 | 87.26 | 1.56 | - | 8 | 133 |
| ≥ 13.8 | 99.29 | 84.77 | 90.33 | 4.86 | 0.002 | 13 | 131 |
| ≥ 16.5 | 98.46 | 90.00 | 93.23 | 6.52 | 0.008 | 0 | 1 |
| **≥ 17.9** | **98.46** | **90.04** | **93.26** | **9.84** | **0.02** | **367** | **199** |
| ≥ 18.5 | 74.87 | 97.97 | 89.13 | 9.88 | 0.02 | 110 | 9 |
| ≥ 20.0 | 67.8 | 98.33 | 86.64 | 36.83 | 0.26 | 175 | 14 |
| ≥ 21.7 | 56.56 | 98.88 | 82.68 | 40.50 | 0.33 | 12 | 11 |
| ≥ 23.5 | 55.78 | 99.32 | 82.66 | 50.68 | 0.44 | 73 | 13 |
| ≥ 29.7 | 51.09 | 99.84 | 81.18 | 82.33 | 0.44 | 795 | 4 |
| > 29.7 | 0 | 100.00 | 61.72 | 320.48 | 0.49 | - | - |

**Legend:** **S%**: sensitivity value expressed as a proportion; **E%**: specificity value expressed as a proportion; **Ac%**: accuracy value expressed as a proportion; **LR+**: positive likelihood ratio; **LR-**: negative likelihood ratio; **TP**: true pairs; **FP**: false pairs.

**Figure 2.**
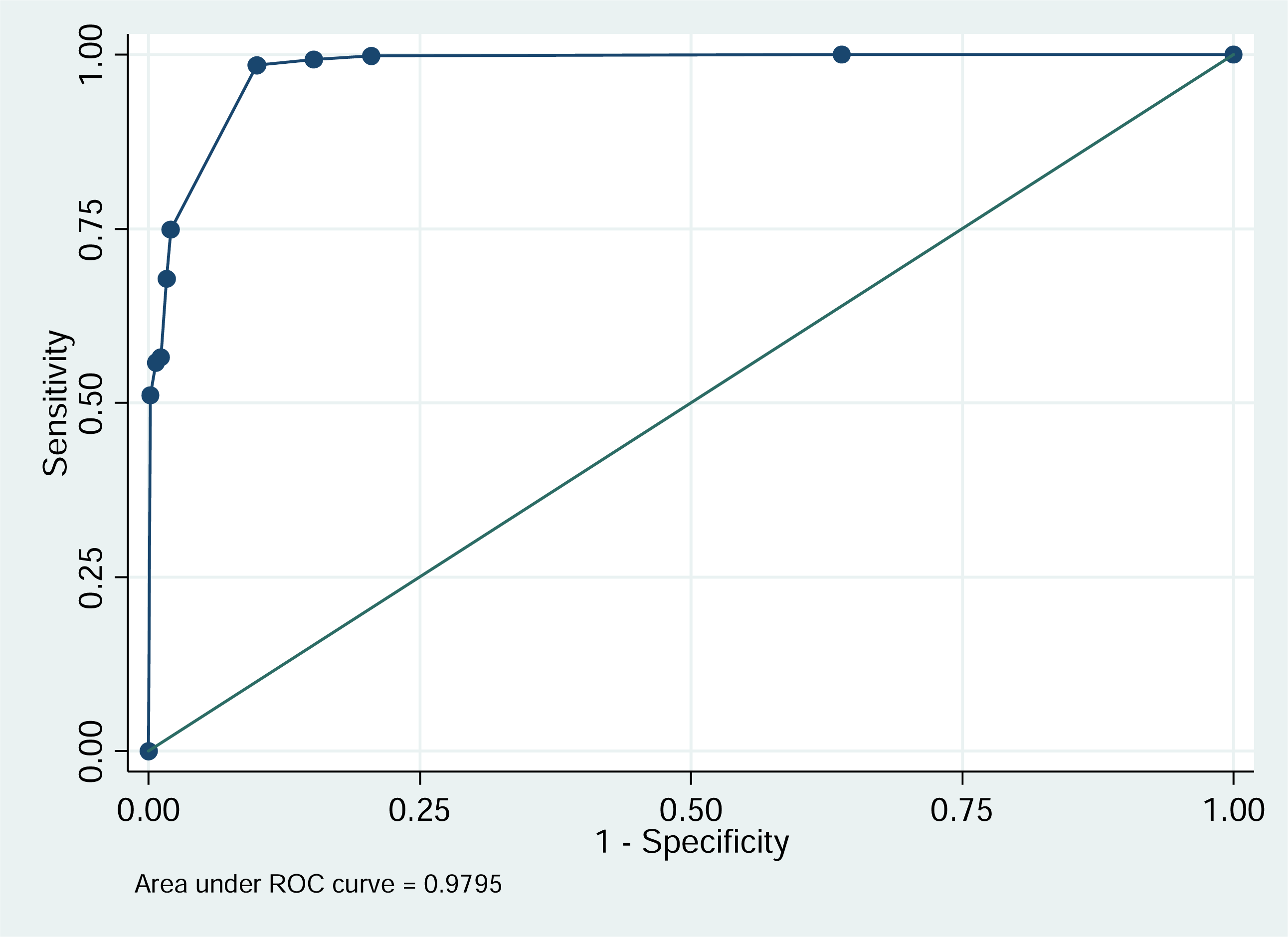
ROC curve of the best blocking and matching strategy (strategy number 1). State of Rio de Janeiro, Brazil, 2015-2020 (n=4065).

**Table 3** presents the characteristics of the study population; the crude mortality rates for the two cutoff points of the scores generated in the probabilistic relationship of the data; and the kappa statistic for each population subgroup. The study population was mostly composed of women (76.72%), whether cisgender or transgender, aged up to 39 years (73.74%), black (65.5%), single (54.4%), and having more than 8 years of school (66.2%). A total of 2.31% of people with disabilities were identified, 4.77% of the population reported some mental or behavioral disorders, and 3.77% had chronic episodes of violence. Most patients (78.12%) had been referred to the assistance network.

**Table 3.** Characteristics of the study population, crude mortality rates (per 1000 persons), and agreement among the cutoff points of the best strategy (#1) for each population subgroup. State of Rio de Janeiro, Brazil, 2015-2020 (n=80.178)

| Variables | n | % | Cutoff $\geq 10.60$ | | Cutoff $\geq 17.90$ | | % Agreement | Kappa | p value |
| --- | --- | --- | --- | --- | --- | --- | --- | --- | --- |
|  |  |  | Deaths | MR | Deaths | MR |  |  |  |
| **Overall** | **80178** | - | 1556 | 19.41 | 1532 | 19.11 | 99.97 | 0.992 | <0.001 |
| **Groups** | **80169** | - | - | - | - | - | - | - | - |
| HCW | 28987 | 36.2 | 285 | 9.83 | 281 | 9.69 | 99.99 | 0.993 | <0.001 |
| WSW | 1159 | 1.45 | 13 | 11.22 | 13 | 11.22 | 100 | 1 | <0.001 |
| CWUnknow | 30884 | 38.5 | 474 | 15.35 | 467 | 15.12 | 99.98 | 0.992 | <0.001 |
| TGW | 472 | 0.59 | 7 | 14.83 | 7 | 14.83 | 100 | 1 | <0.001 |
| MHC | 4496 | 5.61 | 172 | 38.26 | 171 | 38.03 | 99.98 | 0.997 | <0.001 |
| MSM | 534 | 0.67 | 11 | 20.60 | 10 | 18.73 | 99.81 | 0.951 | <0.001 |
| CMUnknow | 13550 | 16.9 | 591 | 43.62 | 580 | 42.80 | 99.92 | 0.99 | <0.001 |
| TGM | 87 | 0.11 | 3 | 34.48 | 3 | 34.48 | 100 | 1 | <0.001 |
| **Age group** | **80178** | - | - | - | - | - | - | - | - |
| 19-29 years | 35327 | 44.1 | 494 | 13.98 | 483 | 13.67 | 99.97 | 0.989 | <0.001 |
| 30-39 years | 23800 | 29.7 | 360 | 15.13 | 353 | 14.83 | 99.97 | 0.99 | <0.001 |
| 40-49 years | 13848 | 17.3 | 342 | 24.70 | 339 | 24.48 | 99.98 | 0.996 | <0.001 |
| $\geq 50$ years | 7203 | 8.98 | 360 | 49.98 | 357 | 49.56 | 99.96 | 0.996 | <0.001 |
| **Race/color** | **62620** | - | - | - | - | - | - | - | - |
| Nonblack | 21629 | 34.5 | 384 | 17.75 | 377 | 17.43 | 99.97 | 0.991 | <0.001 |
| Black | 40991 | 65.5 | 818 | 19.96 | 808 | 19.71 | 99.98 | 0.994 | <0.001 |
| **Marital status** | **47110** | - | - | - | - | - | - | - | - |
| Married and others status | 21461 | 45.6 | 347 | 16.17 | 340 | 15.84 | 99.97 | 0.99 | <0.001 |
| Single | 25649 | 54.4 | 404 | 15.75 | 399 | 15.56 | 99.98 | 0.994 | <0.001 |
| **Schooling** | **29326** | - | - | - | - | - | - | - | - |
| $\geq 8$ years of study | 19423 | 66.2 | 195 | 10.04 | 192 | 9.89 | 99.98 | 0.9896 | <0.001 |
| Up to 8 years of study | 9903 | 33.8 | 235 | 23.73 | 231 | 23.33 | 99.96 | 0.9912 | <0.001 |
| **Persons with disabilities** | **80178** | - | - | - | - | - | - | - | - |
| No | 78325 | 97.7 | 1463 | 18.68 | 1440 | 18.38 | 99.97 | 0.992 | <0.001 |
| Yes | 1853 | 2.31 | 93 | 50.19 | 92 | 49.65 | 99.95 | 0.994 | <0.001 |
| **Persons with a mental or behavioral disorder** | **80178** | - | - | - | - | - | - | - | - |
| No | 76350 | 95.2 | 1368 | 17.92 | 1346 | 17.63 | 99.97 | 0.992 | <0.001 |
| Yes | 3828 | 4.77 | 188 | 49.11 | 186 | 48.59 | 99.95 | 0.994 | <0.001 |
| **Chronicity of violent episodes** | **80178** | - | - | - | - | - | - | - | - |
| No | 77159 | 96.2 | 1493 | 19.35 | 1469 | 19.04 | 99.97 | 0.992 | <0.001 |
| Yes | 3019 | 3.77 | 63 | 20.87 | 63 | 20.87 | 100 | 1 | <0.001 |
| **Referral to the assistance network** | **80178** | - | - | - | - | - | - | - | - |
| No | 17545 | 21.9 | 438 | 24.96 | 428 | 24.39 | 99.94 | 0.988 | <0.001 |
| Yes | 62633 | 78.1 | 1118 | 17.85 | 1104 | 17.63 | 99.98 | 0.994 | <0.001 |

**Legend:** MR: Mortality Ratio; HCW - Heterosexual cisgender women; WSW - Woman who had sex with women; CWUnknow - Cisgender women of unknown sexual orientation; TGW - *Transvestites* and Transgender women; HCM - Heterosexual cisgender men; MSM - Men who had sex with men; CMUnknow - Cisgender men of unknown sexual orientation; TGM - Transgender men

Compared with those in the female population, mortality rates were ∼3 times greater in the male population. Notably, after excluding groups of women with an unknown sexual orientation, the women who had sex with women and transvestites and transwomen had higher mortality rates than did cisgender women. On the other hand, this behavior was not observed in the male group. Higher mortality rates are also observed with advancing age; in black, married and nonsingle people; with less schooling; in people with disabilities or mental and behavioral disorders; and in cases of chronic violence. In contrast, people who received referrals to the health care network had lower mortality rates. With the kappa statistic indicating full agreement for all covariates (p<0.001), the adoption of the cutoff point of ≥17.9 did not produce substantial differences in the interpretation of the results.

## Discussion

The main results of this study are focused on demonstrating the feasibility and good accuracy of determining the probabilistic relationship of data between the SINAN database of interpersonal and self-inflicted violence and the SIM database to obtain information on sexual orientation and gender identity. The study demonstrated the analytical limits of mortality as a function of these variables. Keeping a certain pioneering spirit, as the data linkage technique is used more frequently in health conditions without taking into account the markers of gender and sexuality, these results significantly contribute to overcoming important gaps on the subject in Brazil and in the world. The determination of a linkage strategy, with parsimonious parameters for the identification of true and false pairs, tends to contribute to the use of the technique in the daily routine of health services and in academic production on this topic, which is still considered a scarce area of knowledge.

By adopting preprocessing routines and considering the relationship time as a function of the number of true pairs detected, the recognition of the best blocking and processing strategies constitutes an advance for the systematic adoption of this method in the daily life of the services, especially when considering sexual orientation and gender identity as exposure variables in future analyses. These procedures resulted in a true-pair detection rate with proportions compatible with those found in other investigations whose outcome variable was the registration of death in SIM (21,35).

On the other hand, it is important to note that we are faced with slightly different connecting keys between these investigations. In this article, the linking keys were restricted to the variables “name”, “birthdate”, and “mother’s name”. Although the municipality of residence is traditionally used in probabilistic relationships, in the specific case of our study, this variable could bring more elements of uncertainty than solutions. This is because when reaching groups such as transvestites and transwomen, as well as people known to be victims of violence, depending on the population base used, it is imperative to consider the possibility of constant changes in addresses over time due to stigma and the structure of violence itself that surrounds them (36). There has been a consensus in Brazilian production that housing instability is a factor present in the lives of a significant portion of transvestites and transwomen (11,37). These distinctions in the probabilistic data matching strategies necessitate caution when comparing the data linkage results of different studies.

Complementing this analysis, the definition of a cutoff point in the scores of the probabilistic relationship of data constitutes another important aspect for the advancement of the adoption of this technique in analyses both by health surveillance services and by the academic sector. Recognizing the cutoff point, the manual peer review phase tends to be more efficient (5,25), significantly reducing processing time and improving accuracy in future data relationships. The adoption of scores ≥17.9 reduced 56.16% of the pairs to be reviewed, with 90.04% of the pairs being truly negative. In other words, with high specificity (>90.00%) and with the loss of only 25 true pairs erroneously classified as false (98.46% sensitivity), our strategy 1 demonstrated excellent properties and elements compatible with other linkage strategies with Brazilian datasets (21,35). In addition, when incorporating the deaths into the SINAN database, the degrees of agreement for the kappa indicator are high and close to 1 for all the variables of interest in this study, revealing that the adoption of a higher cutoff point (≥17.9) does not imply losses for future analyses.

The mortality rates observed at the two cutoff points established by the linkage strategy revealed a scenario consistent with the investigations on the subject. That is, advanced age, black race/color, a shorter length of schooling, and disabilities and mental and behavioral disorders (38–40) were associated with higher mortality rates. Thus, these results corroborate the effectiveness of the probabilistic data relationship strategy in the production of reliable information on this topic.

The same occurs in relation to gender markers and sexual orientation. It was observed that transwomen and those who have sex with other women have higher mortality rates than heterosexual cisgender women. Men, whether they are cisgender or transgender, as well as men who have sex with men, also have a higher risk of dying than heterosexual cisgender women (41–44). Thus, having established the cutoff point and the best strategies for capturing information in large datasets, it is essential to scrutinize in more detail how sexual and gender markers affect the risk of dying in the population.

Furthermore, the number of deaths among *travestis* and transgender people is likely higher than that captured in the linkage process, resulting in classification bias. While cisgender men and women have their names established passively, that is, during birth (45), transgender and nonbinary people are actively involved in the choice of this name and still face important bureaucratic barriers to be able to make it compatible with their identity (17). Thus, when considering death as a time-dependent variable, it is possible that the name registered in the notification form, the name of the civil registry (name attributed to birth) (20), can be modified until the moment of death. Thus, in these cases, the death will not be attributed to the person, resulting in a false-negative result and, once again, approaching the risks between the cisgender and transgender people.

Additionally, although the use of the method is promising for the analysis of mortality according to sexual orientation and gender identity, other precautions are necessary. Although the quality of filling out the SINAN has improved over the years (46), notably, the present study found that sexual orientation was not reported (i.e., it was considered “unknown” or “omitted”) in 44.45% of the patients. A similar situation can be observed regarding “race/color”, “marital status”, and, even more markedly, regarding the variable “educational level”.

The quality and completeness of information in Brazilian health information systems have been constant and long-standing challenges for researchers and health managers since the omission of such data greatly compromises the analytical capacity and formulation of public policies (47–49). Because they are constant variables and are known to be associated with asymmetries and inequities in health (50–54), their omission in analytical models, either as a confounding variable or, in some cases, as an exposure variable, can produce strong biases in the production of knowledge on the subject. In this vein, reflecting on professional awareness strategies for the incorporation of these practices of sensitive and attentive questioning to users, as well as awareness that these factors are preponderant for the construction of health actions, are currently urgent elements.

Special care should also be taken concerning the classification of the variable “gender identity”. Although it includes all the responses, the method for classifying the cisgender population in SINAN needs to be closely related since the form used in the system only takes into account the records of *travestis*, trans women and trans men (20). In other words, people who were not registered in the form, or rather, who had an “unknown” record (represented in the form by the options “unknown” or “omitted”), were deemed cisgender. On the other hand, people without a cisgender personality who had not reported their true identity or were not even asked about it, as well as people without a binary personality whose identity could not be recorded, also had an “unknown” record. This scenario especially lowers the sensitivity of the instrument, so in mortality analyses, it is possible that the cisgender data are skewed by miscategorized deaths of others, resulting in a potentially erroneous approximation of the risks among cisgender people and transvestites, transwomen, and transmen (18).

In addition to the limits already exposed and that must be taken into account when interpreting probabilistic relationships with these datasets, an important selection bias must be considered in future analyses. When studying the only database that contains information on sexual orientation and gender identity, the SINAN database of interpersonal and self-inflicted violence, we partially apprehended the actual information experienced by the population. This is because not all people suffer violence, and not all people who suffer violence are notified in health services.

Studies have shown substantial differences in the prevalence of violence between primary surveys and the results obtained by SINAN (55–57). A notable example of these differences is the fact that the surveys indicate psychological violence as the most prevalent (58), while the SINAN data indicate physical violence as the one with the greatest magnitude of violent events (46). This difference may be associated with health professionals’ own understanding of what the phenomenon of violence is, as well as the interpretation of which violence is worthy of reporting and which is not, since notions about this phenomenon are influenced by the sociohistorical conformations of society (59). In other words, culture, values, and social structure directly affect people’s perceptions of what is and is not considered violence. They also produce a kind of stratification of which types of violence are more violent than others and, consequently, those that are worthy of immediate reporting and those that do not need to be recorded urgently (60). Thus, physical and sexual violence, as well as cases that imminently threaten life, might be more common than others. This information bias results in the generation of SINAN results that are different from the reality estimated by surveys. In this context, analyses derived from linkages whose population base is based on SINAN should be interpreted with caution, as the records refer to a population subjected to specific contexts that may be different from those observed in the general population.

This bias can be circumvented by introducing a unique identification key (17) in information systems, such as civil registry numbers, the natural persons registry, or the national registry of the health system, as well as the introduction of the variables “sexual orientation” and “gender identity” into other data collection systems and not only in the SINAN database of interpersonal and self-inflicted violence (17,61). Until then, even with recognized inaccuracies and analytical limits, the performance of procedures involving probabilistic data relationships should be encouraged to better understand the needs of certain subpopulations, improve the technique for capturing information about the LGBT+ population, and thereby advance the formulation of public policies compatible with the needs of this group.

## CONCLUSION

Despite the inaccuracies produced by the data collection format of the variables “gender identity” and “sexual orientation”, the technique of probabilistic data linkage is an important technique with high applicability in the daily routine of health services. With its identification of possible blocking and pairing strategies, as well as the detection of the best cutoff point for linkage scores, this technique becomes more useful in daily research and monitoring by health surveillance services. Although it was not the focus of this investigation, it is notable that women who have sex with women, transvestites, and transgender women have higher mortality rates than do cisgender and heterosexual women. Similarly, people with a male identity have higher mortality rates than women, with no substantial difference according to sexual orientation or gender identity.

## Data Availability

All data produced in the present work are contained in the manuscript

## REFERENCES

1. Baqui P, Bica I, Marra V, Ercole A, van der Schaar M. Ethnic and regional variations in hospital mortality from COVID-19 in Brazil: a cross-sectional observational study. Lancet Glob Health [Internet]. 2020;8(8):e1018–26. Available from: 10.1016/S2214-109X(20)30285-0

2. Ramos D, da Silva NB, Ichihara MY, Fiaccone RL, Almeida D, Sena S, et al. Conditional cash transfer program and child mortality: A cross-sectional analysis nested within the 100 Million Brazilian Cohort. PLoS Med. 2021 Sep 28;18(9):e1003509.

3. Baqui P, Bica I, Marra V, Ercole A, van der Schaar M. Ethnic and regional variations in hospital mortality from COVID-19 in Brazil: a cross-sectional observational study. Lancet Glob Health [Internet]. 2020;8(8):e1018–26. Available from: 10.1016/S2214-109X(20)30285-0

4. Bollyky TJ, Templin T, Cohen M, Schoder D, Dieleman JL, Wigley S. The relationships between democratic experience, adult health, and cause-specific mortality in 170 countries between 1980 and 2016: an observational analysis. The Lancet. 2019 Apr;393(10181):1628–40.

5. Rebora P, Scirè CA, Occhino G, Bortolan F, Leoni O, Cideni F, et al. Development and validation of an electronic database-based frailty index to predict mortality and hospitalization in a population-based study of adults with SARS-CoV-2. Front Med (Lausanne). 2023 May 12;10.

6. Phyo AZZ, Freak-Poli R, Craig H, Gasevic D, Stocks NP, Gonzalez-Chica DA, et al. Quality of life and mortality in the general population: a systematic review and meta-analysis. BMC Public Health. 2020 Dec 6;20(1):1596.

7. Batty GD, Kivimäki M, Frank P. State care in childhood and adult mortality: a systematic review and meta-analysis of prospective cohort studies. Lancet Public Health. 2022 Jun;7(6):e504–14.

8. Ministério da Saúde. Declaração de ÓbitoL: manual de instruções para preenchimento. Brasília: Ministério da Saúde; 2022.

9. Morais RM de, Costa AL. Uma avaliação do Sistema de Informações sobre Mortalidade. Saúde em Debate. 2017 Mar;41(spe):101–17.

10. Ranzani OT, Marinho M de F, Bierrenbach AL. Utilidade do Sistema de Informação Hospitalar na vigilância da mortalidade materna no Brasil. Revista Brasileira de Epidemiologia. 2023;26.

11. Rafael R de MR, Jalil EM, Luz PM, de Castro CRV, Wilson EC, Monteiro L, et al. Prevalence and factors associated with suicidal behavior among trans women in Rio de Janeiro, Brazil. PLoS One. 2021 Oct 22;16(10):e0259074.

12. Montenegro L, Velasque L, LeGrand S, Whetten K, Rafael R de MR, Malta M. Public Health, HIV Care and Prevention, Human Rights and Democracy at a Crossroad in Brazil. AIDS Behav. 2020 Jan 22;24(1):1–4.

13. Spittlehouse JK, Boden JM, Horwood LJ. Sexual orientation and mental health over the life course in a birth cohort. Psychol Med. 2020 Jun 13;50(8):1348–55.

14. Semlyen J, King M, Varney J, Hagger-Johnson G. Sexual orientation and symptoms of common mental disorder or low wellbeing: combined meta-analysis of 12 UK population health surveys. BMC Psychiatry. 2016 Dec 24;16(1):67.

15. Bränström R, Narusyte J, Svedberg P. Sexual-orientation differences in risk of health-related impaired ability to work and to remain in the paid workforce: a prospective population-based twin study. BMC Public Health. 2023 Mar 8;23(1):454.

16. Breda KML, Carava-Morera JA, Rafael R de MRR. Trans trafficking and sex work in Brazil, Costa Rica, and the USA. In: de Chesnay M, Sabella D, editors. Human Trafficking: A Global Health Emergency. Cham: Springer International Publishing; 2023. p. 241–65.

17. Rafael R de MR, Santos HG de S, Caravaca-Morera JA, Wilson EC, Breda KL. Inclusão ou ilusão da identidade de gênero no país com o maior número de assassinatos de transgêneros: um ensaio crítico brasileiro. Escola Anna Nery. 2023;27.

18. Rafael R de MR, Gil AC, Santos HG de S, Caravaca-Morera JA, Breda KL. Ensaio teórico-metodológico sobre validade da informação da identidade de gênero no monitoramento epidemiológico da violência. Revista da Escola de Enfermagem da USP. 2023;57.

19. Benevides BG, Nogueira SNB. Dossiê dos Assassinatos e da violência contra travestis e transexuais no Brasil em 2018. Brasília: Associação Nacional de Travestis e Transexuais no Brasil (ANTRA); 2020. 1–60 p.

20. Ministério da Saúde. Viva: instrutivo notificação de violência interpessoal e autoprovocada. 2nd ed. Brasília: Ministério da Saúde; 2016.

21. Oliveira GP de, Bierrenbach AL de S, Camargo Júnior KR de, Coeli CM, Pinheiro RS. Accuracy of probabilistic and deterministic record linkage: the case of tuberculosis. Rev Saude Publica. 2016;50(0).

22. Machado JP, Silveira DP da, Santos IS, Piovesan MF, Albuquerque C. Aplicação da metodologia de relacionamento probabilístico de base de dados para a identificação de óbitos em estudos epidemiológicos. Revista Brasileira de Epidemiologia. 2008 Mar;11(1):43–54.

23. Quezada-Sánchez AD, Espín-Arellano I, Morales-Carmona E, Molina-Vélez D, Palacio-Mejía LS, González-González EL, et al. Implementation and validation of a probabilistic linkage method for population databases without identification variables. Heliyon. 2022 Dec;8(12):e12311.

24. Asher J, Resnick D, Brite J, Brackbill R, Cone J. An Introduction to Probabilistic Record Linkage with a Focus on Linkage Processing for WTC Registries. Int J Environ Res Public Health. 2020 Sep 22;17(18):6937.

25. Peres SV, Latorre M do RD de O, Michels FAS, Tanaka LF, Coeli CM, Almeida MF de. Determinação de um ponto de corte para a identificação de pares verdadeiros pelo método probabilístico de linkage de base de dados. Cad Saude Colet. 2014 Dec;22(4):428–36.

26. Prindle J, Suthar H, Putnam-Hornstein E. An open-source probabilistic record linkage process for records with family-level information: Simulation study and applied analysis. PLoS One. 2023 Oct 20;18(10):e0291581.

27. Cohen JF, Korevaar DA, Altman DG, Bruns DE, Gatsonis CA, Hooft L, et al. STARD 2015 guidelines for reporting diagnostic accuracy studies: explanation and elaboration. BMJ Open. 2016 Nov 14;6(11):e012799.

28. Silva JC da, Rafael R de MR, Alcântara DC, Carvalho CMSM de, Porphirio MFC, Porphirio MCC, et al. “I can’t breathe”: The effect of intersectionality on access to COVID-19 diagnostic tests in Brazil. IPJS. 2023;9(2):26–36.

29. Centers for Disease Control and Prevention (CDC). National Program of Cancer Registries. Link Plus 3.0 version [Internet]. Atlanta (USA): CDC; 2007 [cited 2023 Jan 4]. Available from: https://www.cdc.gov/cancer/npcr/tools/registryplus/lp.htm

30. Silva DRM e, Luizaga CT de M, Toporcov TN, Algranti E. Concordância e validade dos diagnósticos de cânceres associados ao asbesto no sistema de informação hospitalar do Sistema Único de Saúde. Revista Brasileira de Epidemiologia. 2021;24.

31. Agranonik M, Jung RO. Qualidade dos sistemas de informações sobre nascidos vivos e sobre mortalidade no Rio Grande do Sul, Brasil, 2000 a 2014. Cien Saude Colet. 2019 May;24(5):1945–58.

32. Ministério da Saúde. Saúde Brasil 2020/2021: uma análise da situação de saúde e da qualidade da informação. Brasília: Ministério da Saúde; 2021.

33. Landis JR, Koch GG. The Measurement of Observer Agreement for Categorical Data. Biometrics. 1977 Mar;33(1):159.

34. Altman DG. Practical Statistics for Medical Research. Chapman and Hall/CRC; 1990.

35. Coeli CM, Saraceni V, Medeiros PM, da Silva Santos HP, Guillen LCT, Alves LGSB, et al. Record linkage under suboptimal conditions for data-intensive evaluation of primary care in Rio de Janeiro, Brazil. BMC Med Inform Decis Mak. 2021 Dec 15;21(1):190.

36. Veroneze RT. Vulnerabilidades das travestis e das mulheres trans no contexto pandêmico. Revista Katálysis. 2022 Aug;25(2):316–25.

37. Grinsztejn B, Jalil EM, Monteiro L, Velasque L, Moreira RI, Garcia ACF, et al. Unveiling of HIV dynamics among transgender women: a respondent-driven sampling study in Rio de Janeiro, Brazil. Lancet HIV. 2017 Apr;4(4):e169–76.

38. Sasson I, Hayward MD. Association Between Educational Attainment and Causes of Death Among White and Black US Adults, 2010-2017. JAMA. 2019 Aug 27;322(8):756.

39. Momen NC, Plana-Ripoll O, Agerbo E, Christensen MK, Iburg KM, Laursen TM, et al. Mortality Associated With Mental Disorders and Comorbid General Medical Conditions. JAMA Psychiatry. 2022 May 1;79(5):444.

40. Ferdows NB, Aranda MP, Baldwin JA, Baghban Ferdows S, Ahluwalia JS, Kumar A. Assessment of Racial Disparities in Mortality Rates Among Older Adults Living in US Rural vs Urban Counties From 1968 to 2016. JAMA Netw Open. 2020 Aug 3;3(8):e2012241.

41. Coelho LE, Torres TS, Jalil EM, Cardoso SW, Moreira RI, Calvet GA, et al. Mortality rates by gender and sexual orientation reveal a disproportionally high mortality among cisgender men of unknown sexual orientation and men who have sex with women in a cohort of people living with HIV in Rio de Janeiro, Brazil. The Brazilian Journal of Infectious Diseases. 2023 Mar;27(2):102740.

42. Jackson SS, Brown J, Pfeiffer RM, Shrewsbury D, O’Callaghan S, Berner AM, et al. Analysis of Mortality Among Transgender and Gender Diverse Adults in England. JAMA Netw Open. 2023 Jan 30;6(1):e2253687.

43. Crimmins EM, Shim H, Zhang YS, Kim JK. Differences between Men and Women in Mortality and the Health Dimensions of the Morbidity Process. Clin Chem. 2019 Jan 1;65(1):135–45.

44. Lenart P, Kuruczova D, Joshi PK, Bienertová-Vašků J. Male mortality rates mirror mortality rates of older females. Sci Rep. 2019 Jul 22;9(1):10589.

45. Mota M, Santana AD da S, Silva LR e, Melo LP de. “Clara, esta sou eu!” Nome, acesso à saúde e sofrimento social entre pessoas transgênero. Interface - Comunicação, Saúde, Educação. 2022;26.

46. Pinto IV, Andrade SS de A, Rodrigues LL, Santos MAS, Marinho MMA, Benício LA, et al. Perfil das notificações de violências em lésbicas, gays, bissexuais, travestis e transexuais registradas no Sistema de Informação de Agravos de Notificação, Brasil, 2015 a 2017. Revista Brasileira de Epidemiologia. 2020;23(suppl 1).

47. Flanagin A, Frey T, Christiansen SL, Bauchner H. The Reporting of Race and Ethnicity in Medical and Science Journals. JAMA. 2021 Mar 16;325(11):1049.

48. Santos RV, Bastos JL, Kaingang JD, Batista LE. Cabem recomendações para usos de “raça” nas publicações em saúde? Um enfático “sim”, inclusive pelas implicações para as práticas antirracistas. Cad Saude Publica. 2022;38(3).

49. Correia LO dos S, Padilha BM, Vasconcelos SML. Métodos para avaliar a completitude dos dados dos sistemas de informação em saúde do Brasil: uma revisão sistemática. Cien Saude Colet. 2014 Nov;19(11):4467–78.

50. Cobo B, Cruz C, Dick PC. Desigualdades de gênero e raciais no acesso e uso dos serviços de atenção primária à saúde no Brasil. Cien Saude Colet. 2021 Sep;26(9):4021–32.

51. Evedove AUD, Dellaroza MS, Carvalho WO, Loch MR. Mudança na situação conjugal e incidência de comportamentos de proteção à saúde em adultos com 40 anos ou mais: estudo VigiCardio (2011-2015). Cad Saude Colet. 2021 Sep;29(3):433–43.

52. Malta DC, Bernal RTI, Lima MG, Silva AG da, Szwarcwald CL, Barros MB de A. Socioeconomic inequalities related to noncommunicable diseases and their limitations: National Health Survey, 2019. Revista Brasileira de Epidemiologia. 2021;24(suppl 2).

53. Castro CMS, Costa MFL, Cesar CC, Neves JAB, Sampaio RF. Influência da escolaridade e das condições de saúde no trabalho remunerado de idosos brasileiros. Cien Saude Colet. 2019 Nov;24(11):4153–62.

54. Alcântara DC, Caravaca-Morera JA, Peixoto EM, Rafael RDMR, De Andrade MDC, Gil AC. Intersectionality and transsexuality in the process of discrimination: an integrative review. Revista Enfermagem UERJ. 2022 Dec 30;30(1):e66665.

55. Leite FMC, Santos DF, Ribeiro LA, Tavares FL, Correa ES, Ribeiro LEP, et al. Análise dos casos de violência interpessoal contra mulheres. Acta Paulista de Enfermagem. 2023 Jan 20;36.

56. Lima VM da F, Stochero L, Azeredo CM, Moraes CL de, Hasselmann MH, Marques ES. Characterization and completeness of notification sheet of violence against the older adults in Niterói-RJ, 2011-2020. Epidemiologia e Serviços de Saúde. 2023;32(1).

57. Silva MMA da, Mascarenhas MDM, Lima CM, Malta DC, Monteiro RA, Freitas MG de, et al. Perfil do Inquérito de Violências e Acidentes em Serviços Sentinela de Urgência e Emergência. Epidemiologia e Serviços de Saúde. 2017 Jan;26(1):183–94.

58. Rafael R de MR, Jalil EM, Velasque L de S, Friedman RK, Ramos M, Cunha CB, et al. Intimate Partner Violence Among Brazilian Trans and Cisgender Women Living with HIV or at HIV Risk During COVID-19 Era: Another Epidemic? Transgend Health. 2023 Sep 23;

59. Krug EG, Dahlberg LL, Mercy JA, Zwi AB, Lozano R. World report on violence and health. Geneva; 2002.

60. Butler J. Precarious Life: The Powers of Mourning and Violence. London: Verso; 2004.

61. Canavese D, Polidoro M, Signorelli MC, Moretti-Pires RO, Parker R, Terto Jr. V. A call for the urgent and definitive inclusion of gender identity and sexual orientation data in the Brazilian health information systems: what can we learn from the monkeypox outbreak? Cien Saude Colet. 2022 Nov;27(11):4191–4.

